# Clinical Validation of Self-Measurement for Anogenital Distance in Women

**DOI:** 10.1101/2025.03.23.25324478

**Authors:** Natalie L. Dinsdale, Jenel Maruk, Aiden Bushell, Bernard J. Crespi

## Abstract

Anogenital distance (AGD), the length from the anus to specific genital landmarks, is a well validated, testosterone sensitive, sexually dimorphic biomarker used in many kinds of medical and evolutionary research in diverse species of mammals, including humans. Current research into the effects of testosterone on women’s reproductive health and disease is motivating increased interest in measuring female AGD. Studies quantifying female AGD typically employ a clinician, such as a gynecologist or nurse, to conduct the measurements. This methodology maximizes accuracy but imposes notable limitations on data collection. All participants submitted self-measurements online and completed a small set of questionnaires, including tests assessing spatial cognition. The accuracy of AGD self-measurements, based on agreement between self- and clinic-measurements, was moderate. Measurement accuracy was predicted by performance on the mental rotation test, such that women who performed better on this test demonstrated greater accuracy in measuring the anus to posterior fourchette distance. We describe ideas for improving the accuracy of the self-measurement technique. Self-measurement of AGD would increase the number and diversity of women represented in studies of reproductive health, reduce research expenses, and expedite research into the effects of prenatal testosterone and endocrine disruption on female health and disease.

## Introduction

Anogenital distance (AGD), the distance from the anus to landmarks on the genitalia, is positively associated with prenatal testosterone exposure, such that males typically have substantially longer AGDs than females, and, within each sex, individuals exposed to relatively high levels of prenatal testosterone have longer AGDs than individuals exposed to less (Thankamony et al., 2016). The relationship between AGD and the prenatal testosterone levels makes AGD a useful biomarker in diverse kinds of research, including studies on endocrine disruption, evolutionary endocrinology, developmental origins of disease, and reproductive health (Zarean et al., 2019; Zamani et al., 2023). AGD is especially useful because, unlike 2D:4D digit ratio (Richards, Browne & Constantinescu, 2021), its effects have been well-validated experimentally in studies of mice, rats and rhesus monkeys (Gandelman et al. 1979; Hotchkiss et al. 2007; Abbott et al. 2008).

Given the relationship between AGD and testosterone, comparably more research has been conducted on male AGD than female AGD. However, the roles of prenatal testosterone in shaping women’s reproductive health and disease risks represent a growing area of research (Crespi & Evans, 2023; Dinsdale & Crespi, 2021; Ruth *et al.,* 2020). For example, among women, shorter AGD has been linked with lower adult serum testosterone (Mira-Escolano *et al*., 2014a; Wu *et al*., 2017), lower antimullerian hormone (Fabregues *et al*., 2018), reduced number of ovarian follicles (Mendiola *et al*., 2012), higher risks of endometriosis (Crespi 2024) and premature ovarian insufficiency (Dural *et al*., 2021; Tan *et al*., 2023), and lower risks of polycystic ovary syndrome (e.g., Wu Zhong 2017). Quantifying AGD in large, representative samples of women would facilitate research on the diverse roles of prenatal testosterone in female reproduction, yet relatively few such studies have been conducted.

Measurements of women’s AGD have typically been conducted by professional health care workers in clinical settings. Receiving AGD measurements thus requires women to visit a clinic, disrobe, and have their genitalia examined, touched, and measured by an unfamiliar person, which may limit some women’s willingness to participate in this kind of research. Performing self-measurements of AGD at home may be a useful alternative, but its degree of accuracy has yet to be evaluated.

We investigated the accuracy of an at-home, self-measurement technique for measuring female AGD, by having a cohort of women engage in both clinical measurement, in a health-care setting, and self-measurement, at home, given a set of instructions and figures (Appendix 1). A larger set of additional women only conducted measurements at home, to provide a larger sample of at-home measurement data. The primary objective of the study was thus to quantify the accuracy of an at-home self-measurement technique for two different female AGD measurements (AGD1 - the anus to the posterior fourchette; AGD2 - the anus to the base of clitoral glans) (Thankamony et al., 2016 Salazar-Martinez et al., 2004; Liu et al., 2014). A secondary objective was to assess which, if any, cognitive features predicted the accuracy of self-measurement.

## Methods

### Participants, recruitment, consent, and honoraria

Women between the ages of 18 and 40 years living in Canada who could read and write in English were eligible to participate in the study. Participants located anywhere in Canada were eligible to participate in Group A (self-measurement and online questionnaire group) whereas participants living in or near Saskatoon were eligible to be part of Group B (the subset of women who completed self-measurements at home and received expert measurements at J.M.’s Saskatoon-based massage and wellness clinic).

We recruited Saskatoon-based participants through posters in health and wellness clinics, cafes, libraries, and on poster boards at the University of Saskatchewan campus. We also employed paid advertising on Meta to invite women meeting the above criteria to participate in the study. Two different ads (one targeted to Saskatoon-based women, and one targeted to women living in Canadian cities) were deployed. The online advertisements described the study’s aims, procedures, and honoraria; interested women were invited to message or email Dr. Dinsdale to enroll. Participants gave informed consent and provided their mailing addresses via an online document sent through email. Women participating in Group A were sent $20 CAD via e-transfer after completing the study and women participating in Group B were sent $50 CAD after completing the study.

### Materials and procedures

Enrolled participants were mailed a package containing detailed instructions and supplies for study participation. For Group A participants, the study package included supplies (a handheld mirror, clear plastic ruler marked with metric scale, and soft tape measure) and a detailed booklet that included verbal and visual instructions of how to measure the two different anogenital distances. Participants were instructed to measure the distance from center of the anus to the fourchette (AGD1; Figure 2a), as well as the distance from the center of the anus to the base of the clitoral glans (AGD2; Figure 2b). Women were instructed to measure AGD1 and AGD2 in centimeters three times each, and to record the average of each measurement to later input into the online survey. To do the measurement, women were guided to lay down with the legs apart, holding the handheld mirror with one hand and the ruler (with 0 cm positioned at the anus) with the other hand. We acknowledged to the women that taking these measurements may be awkward or uncomfortable.

In addition to the AGD measurements, women were also instructed to use the soft tape measure to measure the widest part of their hips and narrowest part of their waist, in cm, and to input these measurements into the online survey. Following the recording of bodily measurements, participants were asked to visit the Survey Monkey link provided in the instruction booklet to input their measurements and to answer a variety of questions, which are described below.

Group B participants received the exact same instructions and supplies as Group A participants, but their booklets included additional instructions for contacting J.M.’s clinic after they had completed the self-measurements and online questionnaire. Group B participants were provided with a telephone number and email address to schedule an appointment at the clinic to have their AGD measurements and handgrip strength assessed by J.M. Participants were encouraged to contact the clinic to book their appointment within two days of completing the at-home portion of the study.

The clinical AGD measurements were conducted by J.M. using the same instructions and supplies as the participants. Participants disrobed from the waist down and laid down on a clean massage table. Each distance (clin-AGD1 & clin-AGD2) was measured three times and both mean distances were recorded by J.M. and securely sent to Dr. Dinsdale. Each participant in Group B also received a handgrip strength assessment. Participants squeezed a dynamometer three times each with the right and left hand and all values were recorded. The handgrip strength and waist and hip measurement data will be used in another project.

### Online questionnaires

The online survey was hosted on Survey Monkey. Participants were instructed to first provide their study identification code (for anonymity) and then confirm that they had read and understood the AGD measurement instructions. After inputting their AGD and body measurements (in cm) into Survey Monkey, participants answered a series of questions, including demographic questions (age, ethnicity, education level), medical-historical questions (height, weight, age at menarche, number of births, medical diagnosis, medications), and a series of health, personality, and cognitive questionnaires. Only data from the cognitive questionnaires are included in the present study.

We selected three tests of cognitive ability that tend to show sex differences and effects from testosterone in the literature. Firstly, we included a 20-item Folk Physics test (Baron-Cohen et al., 2001a), a 20-item mental rotation matching test (Peters et al., 1995); and the 10-item Brief-Reading the Mind in the Eyes Test (RMET-10; Olderbak et al., 2015). The Folk Physics test assesses a subject’s ability to correctly deduce the causes of non-agentic movement, such as the outcomes of a gear turning or pulley lifting. Participants are given a series of images and then asked to select the correct outcome from multiple choice options about what is or what will occur as the depicted gear, pulley, or lever moves. One point is given per correct answer, for a range of 0-20 points.

The matching Mental Rotation Test is a modified version of Peters et al. (1995) test that presents pairs of three-dimensional geometric shapes rotated to different orientations. Participants must choose ‘match’ or ‘no match’ to correctly assess whether the two shapes are identical (‘match’ but rotated to different orientations) or distinct (‘no match’ regardless of their orientation in space). The mental rotation and intuitive physics tests were selected because they both assess spatial abilities (especially mental rotation) and physical science skills (especially intuitive physics), and both show male biases in their scores (Lauer et al. 2019).

The brief Reading the Mind in the Eyes presents participants with a photograph of the eye region of a person’s face. Participants are given multiple choice answers and are instructed to select the correct emotion expressed through the person’s eyes in the photographs. The RMET (Baron-Cohen et al., 2001b), and the RMET-brief, are widely used as tests of social cognitive ability, and show clear female biases in their scores (Kirkland et al. 2013)

### Ethical approval and funding

This study was approved by the Simon Fraser University Research Ethics Board (Study Number: 30001642) and supported by a grant to Dr. Crespi (NSERC Discovery Grant 2019- 04208).

### Statistical analyses

Data was compiled and organized in an Excel spreadsheet from J.M. and Survey Monkey, and all subsequent statistical analyses were conducted in R (version type: R 4.4.1).

## Results

A total of 173 women enrolled in the study and were mailed study packages (Group A: n = 123; Group B: n = 50). Of these enrolled participants, 122 women completed the study tasks (Group A: n = 94; Group B: n = 28). Participants who did not respond ‘yes’ to the first question of the online survey, indicating that they read and understood the self-AGD measurement instructions, were excluded from analyses (n = 1). One participant was excluded as she reported an age of 41 years and the eligible age range was 18-40 years. Further, if participants left the majority of survey questions blank, they were excluded (n = 1).

The final sample included a total of 119 women who completed self-measurements of AGD and the online survey, and a subset of 28 women who completed self-measurements, the online survey, clinical measurements of AGD, and handgrip strength assessments. Table 1 summarizes demographic data for Group A (self-measurements) and Group B (self and clinic measurements).

**Table 1.** Demographic and descriptive data of Group A (self-measurement group) and Group B (self-plus clinic-measurement group)

| **Demographic Feature** | **Group A (n = 91)** | **Group B (n = 28)** |
| --- | --- | --- |
| **Age** | 31.8 years | 31.0 years |
| **Education level** | Median education level was completed college degree | Median education level was completed college diploma |
| **Ethnicity** | 54% European origins<br>21% Asian<br>11% First Nations<br>7% Mixed<br>3% Black<br>2% Hispanic<br>2% Middle Eastern | 79% European origins<br>14% Mixed<br>3.5% First Nations<br>3.5% Black |
| **BMI** | 27.3 | 31.0 |
| **Age at menarche** | 12.4 years | 11.8 years |
| **Ever experienced pregnancy (Y/N)** | 49% yes | 46% yes |
| **Given birth (how many times)** | 23% - one birth<br>9% - two births<br>3% - three births<br>1% - four births<br>1% - five births | 18% - one birth<br>14% - two births |
| **On hormonal birth control (Y/N)** | 22% yes | 18% yes |
| **Sexual orientation** | 59% heterosexual<br>31% bisexual<br>2% homosexual<br>2% asexual<br>5% other | 54% heterosexual<br>32% bisexual<br>7% homosexual<br>7% asexual |

### Descriptive statistics

To assess the agreement between the self- and clinic-measurements of female AGD, we first compiled the descriptive statistics (Table 2) and identified potential covariates (Table 3). The women who received clinic measurements of their AGD are also represented in the self-measurement column of Table 2. One notable difference between the self-AGD measurements and the clinic-AGD measurements is the wider range of self-measurements, such that the range of self-measured AGD1 and AGD2 extends to notably smaller values than do the clinic measurements. Thus, approximately 5-8% of self-measurements are likely inaccurate as they are too short to be biologically plausible (below 20 mm for AGD1 and below 50 mm for AGD2) when compared to ranges of adult female AGDs from previous research (Mendiola et al., 2012).

**Table 2.**
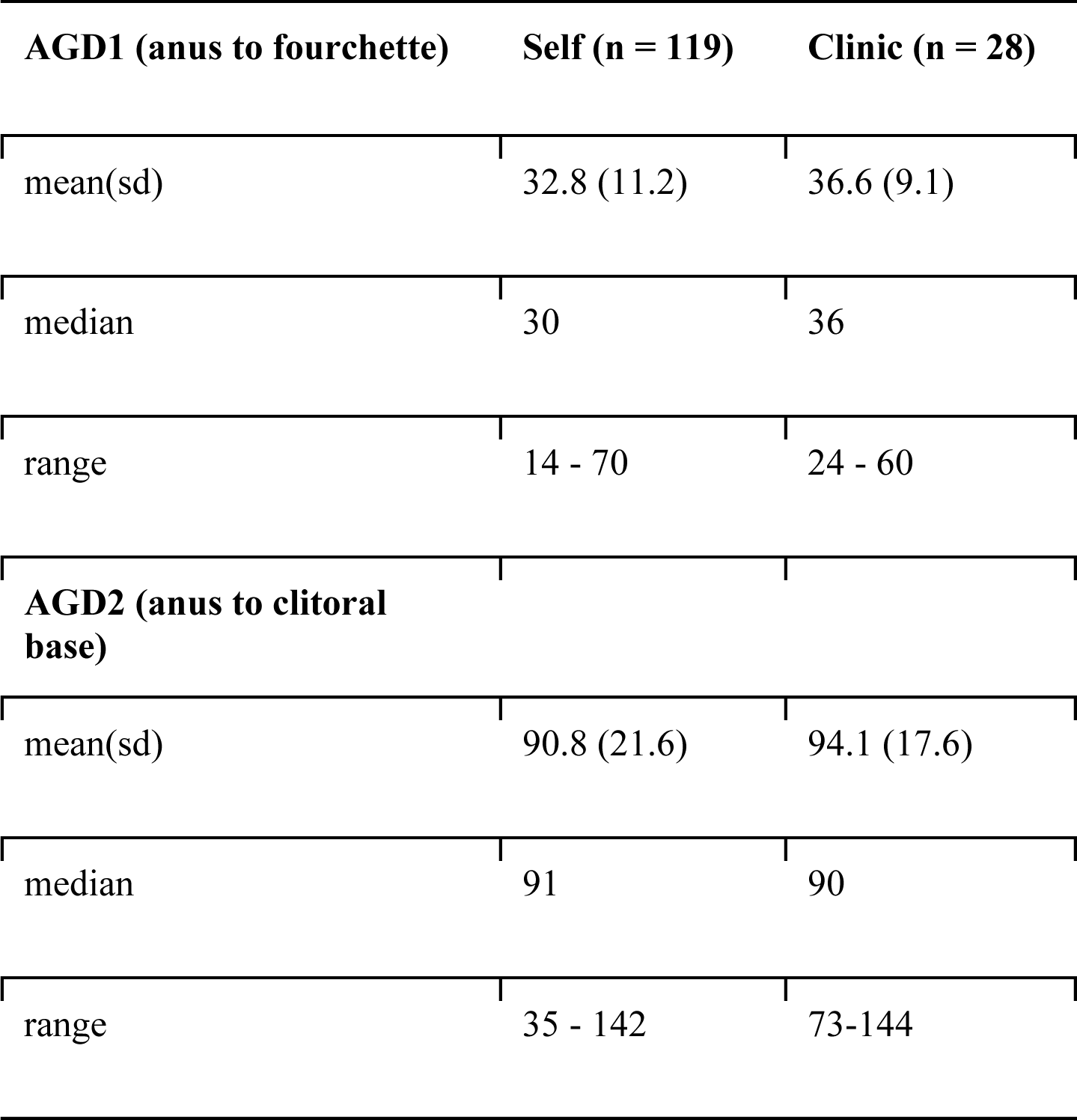
Descriptive statistics of self- and clinic-measurements of AGD1 (anus to fourchette) and AGD2 (anus to base of clitoris) in women who completed self measurements (n = 119) and subset of women who received clinic measurements (n = 28). All measurements were taken in cm to one decimal point, and are converted to mm for easy comparison with other studies.

**Table 3.** Correlation coefficients (rho) between AGD measurements and potential covariates.

| **AGD** | **Height** | **Weight** | **BMI** | **WHR** | **Age** | **Education level** | **Age at menarche** |
| --- | --- | --- | --- | --- | --- | --- | --- |
| **AGD1 (self)** | 0.15 | 0.35*** | 0.27** | 0.19* | 0.04 | -0.10 | -0.08 |
| **AGD2 (self)** | 0.14 | 0.35*** | 0.30** | 0.12 | 0.04 | -0.25** | -0.03 |
| **AGD1 (clin)** | 0.01 | 0.57** | 0.65*** | 0.40* | -0.05 | 0.21 | -0.34◊ |
| **AGD2 (clin)** | -0.18 | 0.45* | 0.50** | 0.31 | -0.03 | 0.06 | -0.11 |

**Significance levels:** ◊ $p < 0.10$ ; \* $p < 0.05$ ; \*\* $p < 0.01$ ; \*\*\* $p < 0.001$

The normality of each of the four distributions (self-AGD1; self-AGD2; clin-AGD1; clin-AGD2) was assessed using visual analysis with QQ plots as well as statistical tests of normality. The four QQ plots indicated some deviations from normality, which were confirmed by a series of Shapiro-Wilks tests. Significant results, indicating non-normality, were returned for self-AGD1 (W = 0.94, p = 0.0001), clin-AGD1 (W = 0.91, p = 0.02), and clin-AGD2 (W = 0.9, p = 0.01). The null hypothesis of normality was not rejected for self-AGD2 (W = 0.99, p = 0.27). Non-parametric tests were selected for subsequent analyses.

Consistent with Mendiola et al. (2012), we found that AGD1 and AGD2 were strongly positively correlated. For self-measured AGD (n = 119), a Spearman rank-order correlation between AGD1 and AGD2 gave rho = 0.53, p<0.0001. For clinic-measured AGD (n = 28), a Spearman rank-order correlation gave rho = 0.57, p<0.01. Such strong correlations are expected, in part, because the AGD1 distance represents a portion of the AGD2 distance.

Next, we examined the effects of potential covariates (height, weight, body mass index, waist-to-hip ratio, age, educational level, age at menarche, and history of pregnancy or birth) on all AGD measurements. Body mass index (BMI) was calculated from self-reported participant height and weight. One outlier was removed from height and BMI analyses (participant reported a very short, implausible height which gave an extreme and unlikely BMI value). Waist-to-hip ratio (WHR) was calculated from self-measured waist and hip circumference. We used Spearman’s rank-order correlation test to conduct these correlation tests. Table 3 shows the correlation coefficients between each of the four AGD measurements and the potential covariates. Consistent with previous research on female AGD (Mendiola et al, 2014), all AGD measurements were significantly and positively associated with weight and BMI.

We used ANOVA to investigate the potential effects of pregnancy (yes or no) and number of births (1-5) on the four AGD measurements. There were no significant effects of pregnancy or births on any AGD measurements (all p values > 0.05). Any significant covariates are considered in relevant, subsequent analyses.

### Objective 1 - Agreement between self-measured and clinic-measured female AGD

We plotted clin-AGD1 against self-AGD1, and clin-AGD2 against self-AGD2, to visually identify any outliers. Each plot indicated one clear outlier. In the AGD1 data, one participant self-measured at a length double to the clinic measurement and in the AGD2 data, one participant self-measured at a length half of the clinic measurement. These outliers were removed for the agreement analyses. We used Spearman’s correlation test and the intraclass correlation test to assess inter-measurement agreement for AGD1 and AGD2.

For AGD1, self-measurements and clinic measurements were significantly and positively correlated with one another (rho = 0.48, p = 0.01, N = 27). For AGD2, self-measurements and clinic measurements were also significantly and positively correlated (rho = 0.44, p = 0.02, N = 27). Figure 1a demonstrates the linear relationship between self-AGD1 and clin-AGD1, and Figure 1b demonstrates the linear relationship between self-AGD2 and clin-AGD2. The regression lines were calculated using the ‘lm’ function in R. Removing the self-measured AGD values that were biologically implausible (AGD1 < 21 mm; AGD2 < 60 mm) modestly increased rho for AGD1 (rho = 0.51, p = 0.007) and did not change rho for AGD2.

**Figure 1a.**
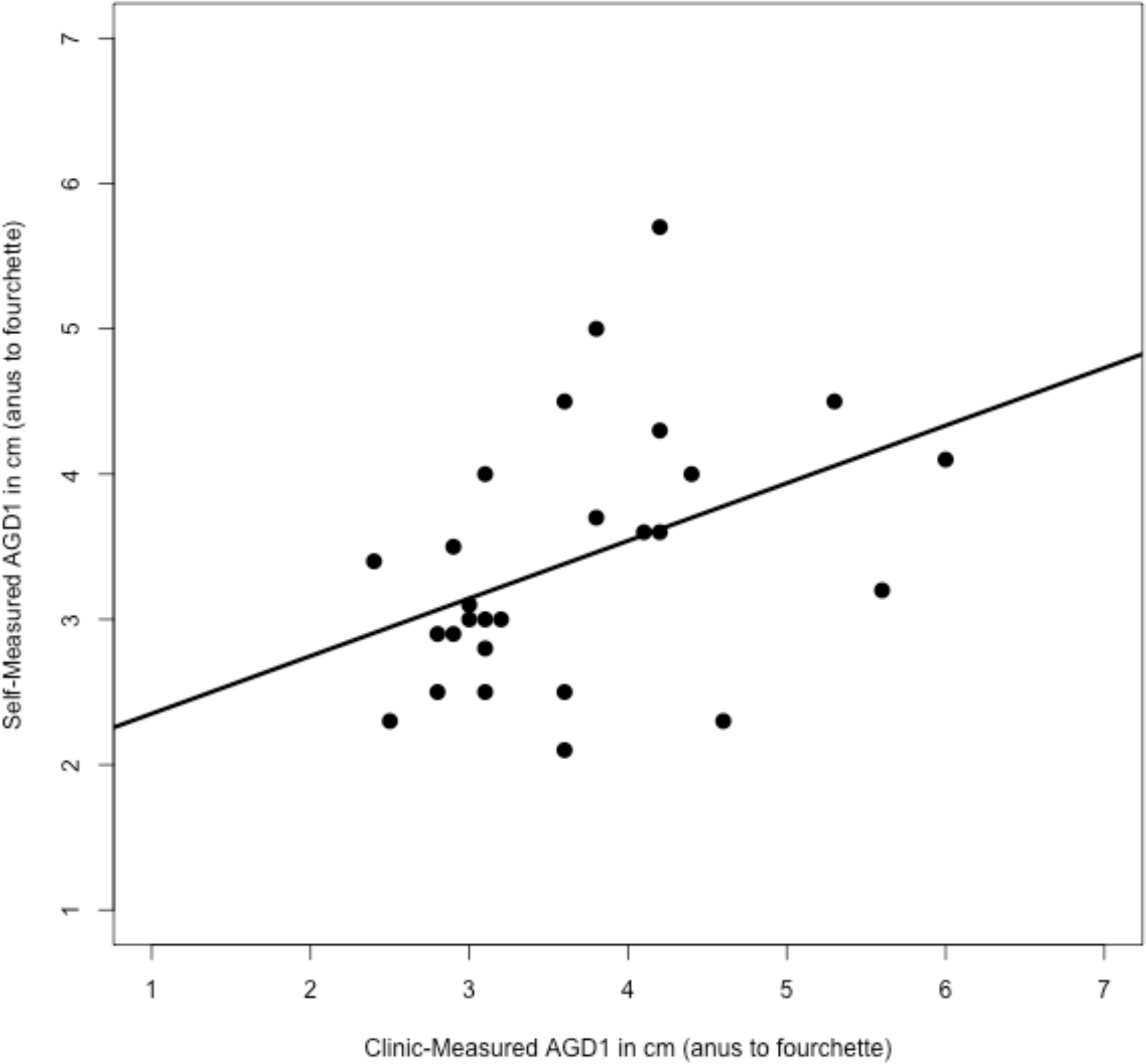
Clinic-measured AGD1 (anus to fourchette) plotted against self-measured AGD1 (anus to fourchette) with regression line.

**Figure 1b.**
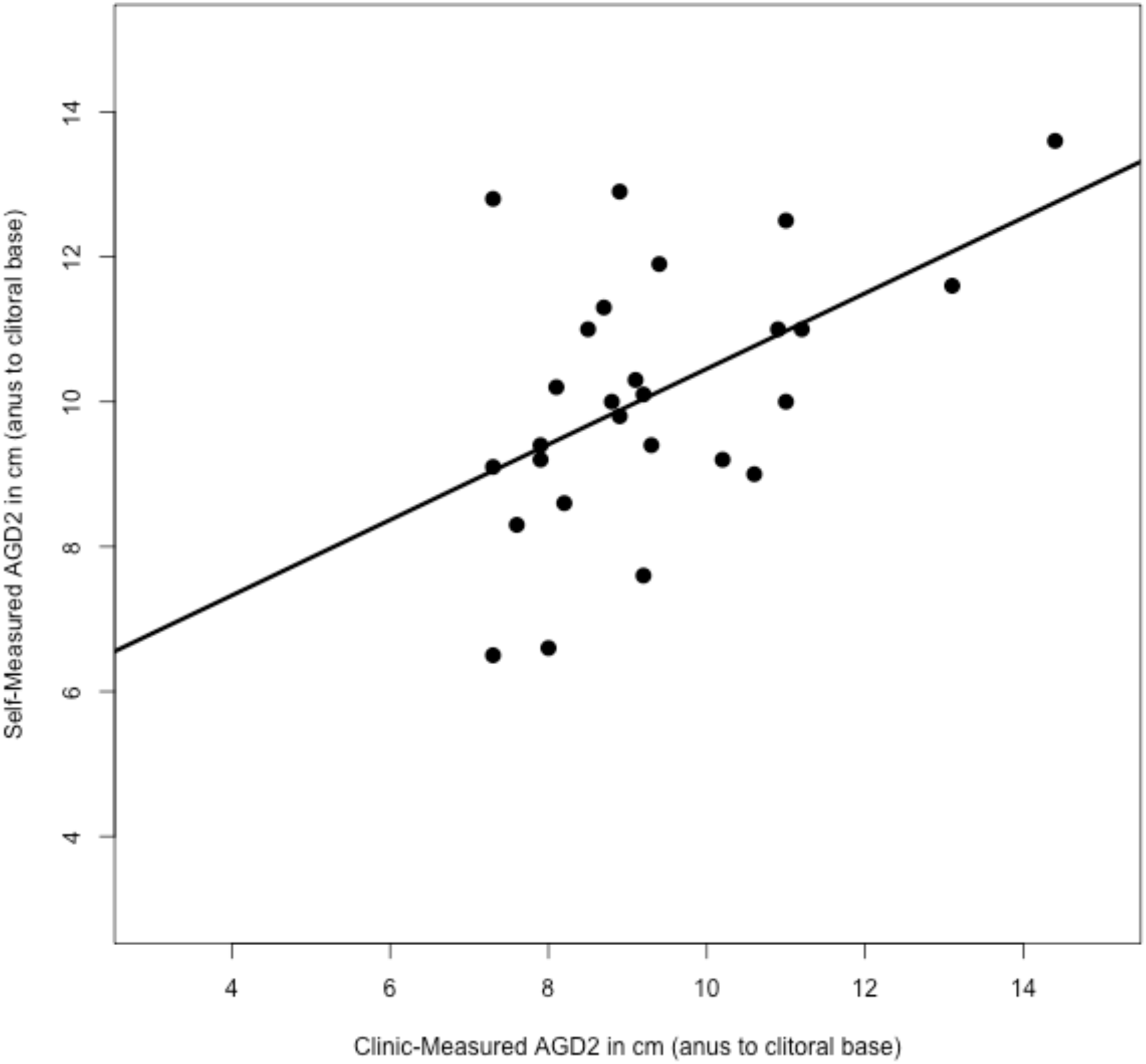
Clinic-measured AGD2 (anus to clitoral base) plotted against self-measured AGD2 (anus to clitoral base) with regression line.

We conducted an intraclass correlation test using the ‘irr’ package in R, after log-transforming the data to adjust for non-normality. This test assesses the reliability among measurements made by multiple judges or raters. For AGD1, the ICC value (n = 27, raters = 2) was 0.42. For AGD2, the ICC value (n = 27, raters = 2) was 0.46. Both of these intraclass correlations are considered moderate in their level of agreement (i.e., agreement between J.M.’s clinic measurements and the subject’s self-measurements).

### Objective 2 - Analyzing predictors of participant self-measurement accuracy

To assess cognitive traits that might impact accuracy of self-measurements, we calculated a linear regression of self-AGD values on clinical-AGD values (for both AGD1 and AGD2) and determined the absolute value of the residuals extracted from the linear regression model. We then tested for correlations between the residuals and the three cognitive tests: mental rotation; intuitive physics; and the RMET. Table 4 summarizes the correlation coefficients and p-values for these six associations. The only significant correlation was between mental rotation performance and AGD1 residuals, indicating that higher mental rotation test scores predicted higher accuracy in self-measuring AGD1.

**Table 4.** Correlations between the absolute values of residuals extracted from lm(clin-AGD1∼self-AGD1) and lm(clin-AGD2∼self-AGD2), and three cognitive test scores (mental rotation; intuitive physics; and RMET).

| **N = 28** | **Mental rotation** | **Intuitive physics** | **RMET** |
| --- | --- | --- | --- |
| **AGD1 residuals** | -0.39* | -0.25 | -0.21 |
| **AGD2 residuals** | -0.082 | -0.24 | -0.26 |

**Significance levels:** $\diamond p < 0.10$ ; \* $p < 0.05$ ; \*\* $p < 0.01$ ; \*\*\* $p < 0.001$

## Discussion

Anogenital distance is a well-validated biomarker of prenatal testosterone in women and men, but its use as a metric has been limited by the invasive methodology required for highly accurate clinical measurements. We have assessed the accuracy of a self-measurement technique for female AGD, as well as evaluating cognitive predictors of self-measurement accuracy. Our main result is that self-measurement is feasible and viable as a means of data collection but would benefit from further refinement of its methodology.

The degree of accuracy of self-measurements of AGD, compared to clinical measurements by a health-care professional, is moderate, as indicated by positive correlations on the order of 0.45-0.50. Correlations for both AGD1 and AGD2 were statistically significant, such that self-measurements provide information about clinical measurements, but with considerable variation. To the extent that such variation is random with regard to the goals of any given study, statistical analyses using self-measured AGD will be conservative, relative to those that use clinical measurement, due to increase measurement error. Consistent with previous work on AGD in women (Mendiola et al., 2012), self-measured AGD1 and AGD2 were also strongly and positively associated and positively related with BMI and weight. These associations also provide support for the accuracy of the technique.

Self-measurements of AGD were, additionally, more accurate (as indicated by residuals from linear regression) among women who scored higher on a test of three-dimensional spatial abilities, the Mental Rotation Test, though not on a test of physical science abilities (the Intuitive Physics test) or a test of cognitive empathy skills (the Reading the Mind in the eyes test). These findings suggest that measurement accuracy depended in part on ability to orient in space, as is required when self-measuring AGD and comparing landmarks on one’s anatomy to the scales of a ruler. Scores on the mental rotation test have been positively associated with three-dimensional measurement abilities in previous work (Bogomolova et al. 2023) and can also be increased through short periods of training (Wang et al. 2023).

Based on these findings, and our experiences during this study, we can suggest several means by which the accuracy of AGD self-measurement may be improved. First, participants might be provided training in mental rotation (e.g., Jost & Jansen, 2021), given that this ability predicted more-accurate AGD measurement. Second, participants may usefully be provided with instruction, in addition to that provided here, regarding the relevant anatomical landmarks, using a minimum of technical language; some subjects were unfamiliar with the relevant anatomy, or its associated names, and line drawings with arrows for indication may have provided them with insufficient information. Third, some participants were apparently not accustomed to the practice of taking measurements on a linear numerical scale; having them practice, and learning about, taking such measurements on diagrams provided on paper, before the actual self-measurement, likely would have also improve accuracy for some individuals. Aside from the medical benefits that could stem from an improved validated protocol for self-measurement of AGD, such procedures may also serve to help better engage women with their own reproductive health and familiarize them better with the practices of science and how to participate in its progress.

## Data Availability

Data will be made available upon reasonable request

## Acknowledgements

The authors thank the subjects for their participation, and NSERC for financial support.

## Declaration of Interest

The authors declare no competing interests.

---

**Supplementary Figure 1.**
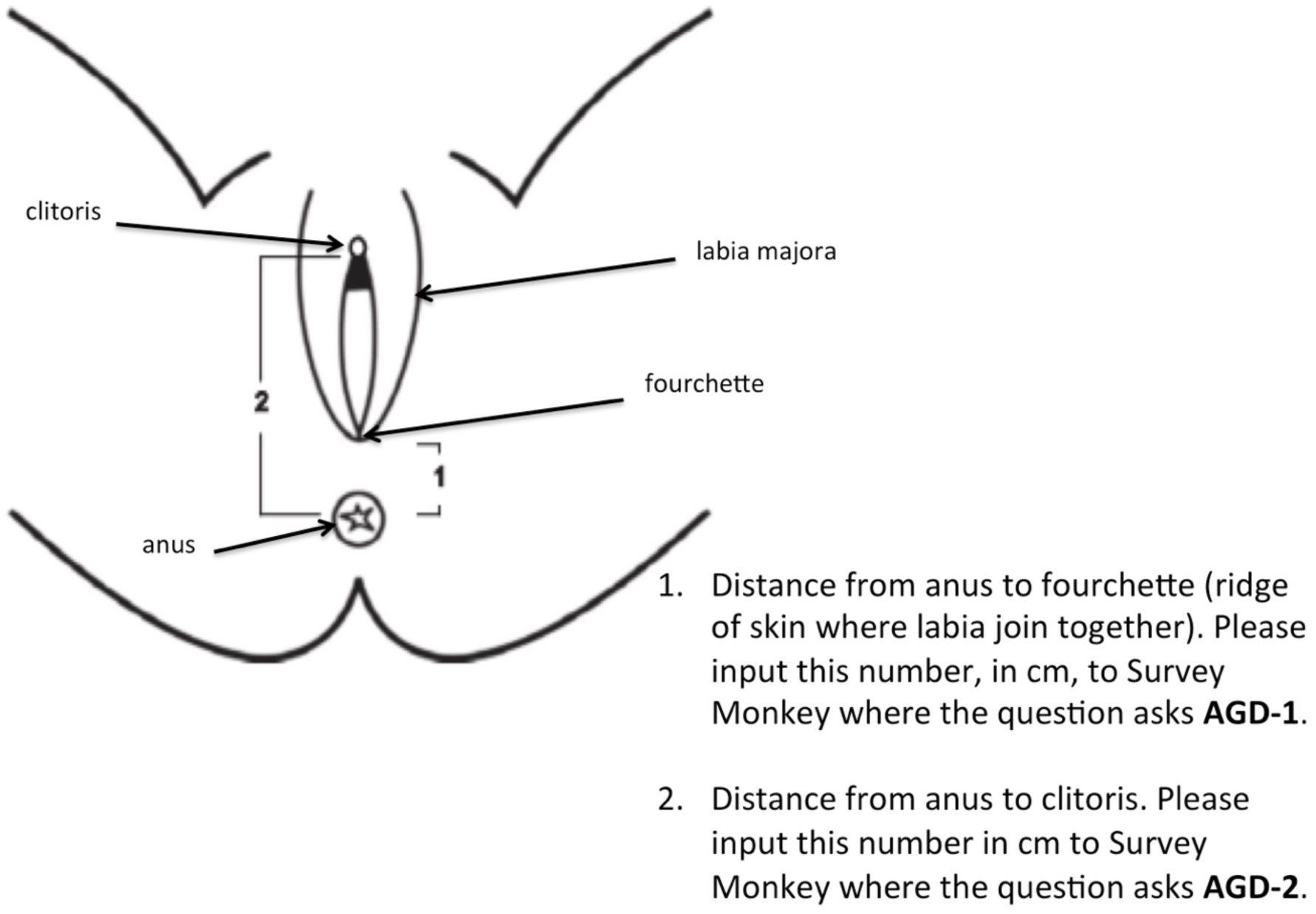
Simplified schematic diagram of genital landmarks used in the calculation of two anogenital distances (AGDs) in women (1 - distance from anus to fourchette; 2 - distance from anus to clitoris). Please note that studies use different parts of the clitoris for measuring female AGD: some studies measure to the anterior surface of clitoris (eg. Priskorn et al., 2017; Mendiola et al. 2012) whereas others measure to the base of the clitoris (eg. Serwah Bonsu Asafo-Agyei et al., 2017; Adekoya et al., 2019). The present diagram and study measured to the base of the clitoris.

## Notes

### Competing Interest Statement

The authors have declared no competing interest.

### Author Declarations

This study was approved by the Simon Fraser University Research Ethics Board Study Number 30001642.

